# An Exploratory Study on the Long-Term Impact of Voiding Cystourethrogram (VCUG)

**DOI:** 10.64898/2026.04.15.26350983

**Authors:** Andrea McDonald, Kathleen Sullivan

## Abstract

**OBJECTIVE:** This study investigates the long-term impacts of childhood exposure to voiding cystourethrogram (VCUG), a diagnostic procedure for vesicoureteral reflux. Primary outcomes include long-term health outcomes, mental health disorders, healthcare avoidance, and participation in risky behaviors compared to a control group.

**METHODS:** A 9-month retrospective cohort study was conducted with adults who received most of their medical care in the U.S. Respondents self-reported health metrics, behaviors, and outcomes via a 20-minute survey. Respondents were divided into two groups: those who remembered undergoing at least one VCUG in childhood (VCUG group), and those who did not (control group).

**RESULTS:** Of 334 respondents, 204 (61%) were in the VCUG group (mean age: 29, 70% female) and 130 (39%) were controls (mean age: 34, 70% female). Notable findings include: 47% of VCUG respondents were diagnosed with depression compared to 27% of controls (χ² = 13.51, p = 0.0002); 15% of female-born VCUG respondents reported they would never visit a gynecologist compared to 2% of controls (χ² = 17.29, p < 0.001); 34% of VCUG respondents smoked regularly compared to 5% of controls (χ² = 35.85, p < 0.001); and 11% of VCUG respondents regularly missed work compared to 1% of controls (χ² = 11.61, p = 0.0006). These findings highlight the need for further research and clinical consideration of VCUG’s long-term consequences.

**CONCLUSIONS:** This study suggests that the effects of childhood VCUG extend into adulthood. Our findings underscore the need to reassess informed consent protocols and consider full-scale studies to minimize bias.

## Introduction

Vesicoureteral reflux (“reflux”) (1) is a common pediatric condition affecting up to 2% of all children (2) in which urine flows backward from the bladder into the ureters and/or kidneys. Reflux presents as, and leads to, recurrent urinary tract infections (3). Childhood reflux frequently resolves on its own (4), but more severe presentations can lead to permanent kidney damage if left untreated (5).

Voiding cystourethrogram (VCUG) is the current gold standard diagnostic test for reflux (1) and has been used for over 60 years (6). Many providers across health disciplines order VCUGs to confirm a diagnosis (1). Despite the procedure’s significance and widespread utilization, there is currently no enforced protocol to order and perform VCUGs in children, with a notable lack of guidelines for patients beyond 2 years of age (1). Publicly available data cannot substantiate how frequently and at what ages VCUGs are performed, though it is estimated that as many as 1 million children in the United States under the age of 18 will receive at least one VCUG per year. A given patient may undergo multiple VCUGs over several years, multiple VCUGs in the same year, or even multiple VCUGs (otherwise known as “voiding cycles”) in the same session (1).

Due to the sensitive nature of the actions required to perform them, VCUGs have been identified as potentially highly traumatic experiences for children. Despite this, the use of sedation is inconsistent because of concerns surrounding its impact on diagnostic outcomes. In 2004, VCUG patients were recruited as proxies for child sexual abuse victims (7) after a physician panel determined that patients who underwent VCUG experienced enough commonalities to molestation that these patient groups could offer valuable information on memory after child sexual abuse (8, 9).

While there is some evidence that VCUG patients have enduring trauma, studies on the topic bear follow-up timelines of no longer than 6 months post-procedure. With these gaps in mind, the long-term goal of this body of work is to elucidate the long-term effects of VCUGs, with a goal of informing current and future approaches to this practice.

## Methods

### OVERVIEW

This was an exploratory study conducted as a retrospective cohort study based on self-reported health information. We obtained approval to conduct this study from the University of Pennsylvania Institutional Review Board. Questions were reviewed by peers with expertise in clinical pediatrics, medical education, consulting, and survey design. The survey was distributed and collected using REDCap (10). To minimize bias, this study was advertised as an exploration of pediatric experiences and adult health, rather than VCUG trauma.

### ELIGIBILITY CRITERIA

Eligible respondents must have been at least 18 years old at the time of the study, able to recall their medical history, and received “most or all” (left to interpretation) of their childhood medical care, including any applicable childhood VCUGs, in the United States. Those who underwent VCUGs or most medical care in a country outside of the United States were not eligible to participate in the study. To be included in the VCUG group, respondents must have had their VCUG during childhood (<18 years old) at an institution in the United States.

Respondents were classified as controls if they did not recall ever undergoing VCUG. Recruitment lasted over eleven months, from October 2022 through September 2023, via a study website, email, LinkedIn, Instagram, Reddit, Facebook, and various independent blogs.

Volunteers also distributed the study flyer to various online and personal channels. Those who reached out were sent an FAQ describing the goals of the study, inclusion criteria, time frame, and next steps. The survey remained open for roughly nine months, from January to September 2023. VCUG respondents were offered $5 compensation in the form of an Amazon gift card, emailed upon survey completion.

### SURVEY DESIGN

Interested parties may step through the study as if they were a participant [here](https://redcap.link/studycontrols). We asked respondents to inform on a variety of potential health and lifestyle factors and outcomes, with a focus on five key areas:

#### Perceptions of physical and mental health

We investigated respondents’ perceptions of their health by querying their overall physical and mental health on a 0 (extremely poor) to 100 (excellent) sliding scale. We phrased this question as, “how would you evaluate your overall [physical or mental] health? Use the sliding scale to rate.” T-tests were employed to discern mean differences between the VCUG and control groups in these self-assessments.

#### Prevalence of long-term illness

We examined the relationship between VCUG exposure and the prevalence of various pelvic and psychological conditions: anxiety, depression, post-traumatic stress disorder (PTSD), obsessive-compulsive disorder (OCD), restless genital syndrome (RGS) / persistent genital arousal disorder (PGAD), hypertonic pelvic floor (HPF), and bipolar disorder (BPD). Respondents who indicated ambiguity about their diagnosis (“I have not been diagnosed, but that sounds like something I might have”) were grouped as “not diagnosed” along with those who confirmed no diagnosis. This allowed us to focus exclusively on confirmed diagnoses (“Yes, I have been diagnosed”). This approach offers a clear delineation of VCUG’s association with diagnosable conditions, while future consideration of ‘maybes’ could provide additional insights into the recognition and diagnosis of these conditions among affected individuals.

Anxiety was characterized as feelings of tension, worried thoughts, and physical changes like increased blood pressure (11). Depression was characterized as feelings of persistent sadness or hopelessness and/or loss of interest in activities, causing impairment in daily life (12). Post-traumatic stress disorder (PTSD) was characterized as a difficulty or failure to recover after experiencing or witnessing a terrifying event, with symptoms such as nightmares, flashbacks, avoidance of situations that bring back memories of the trauma, heightened reactivity to stimuli, anxiety, or depressed mood” (13). Obsessive-compulsive disorder (OCD) was characterized as repetitive and persistent, often unreasonable thoughts and fears (obsessions) that lead to compulsive behaviors (14). Restless genital syndrome (RGS) / persistent genital arousal disorder (PGAD) were characterized as unwanted feelings of excessive and/or persistent genital and/or clitoral stimulation WITHOUT voluntary sexual desire or activity. This can include pain, pressure, swelling, itching, arousal, discomfort, activation, and/or stimulation (15). Hypertonic pelvic floor (HPF) was characterized by muscles in the pelvic floor becoming too tense and unable to relax. Many people experience pelvic health concerns such as constipation, painful sex, urinary urgency/frequent urination without relief, and/or pelvic pain (16). Bipolar disorder (BPD) was characterized as “episodes of mood swings ranging from depressive lows to manic highs” (17).

#### Attitudes towards medical care

We investigated attitudes towards medical care among respondents, focusing on routine physician visits, gynecological care, pap smears, and consultations with urologists. Responses chose between 1) “Yes - I have before (at least once)”, 2) “No - Never have, but I will if I need to”, and 3) “No - Never have, and I won’t (even if I need to)”, enabling us to discern patterns of medical care engagement and refusal. We grouped responses 1 and 2 as “engaged” and response 3 as “refused”, to provide a clear basis for analyzing patient behavior towards adult healthcare utilization post-childhood VCUG.

#### Quality of life factors

We probed the association between VCUG and perceived quality of life in adulthood, focusing on daily interactions and behaviors that reflect personal health perception and interpersonal relations. Respondents evaluated their experiences across six domains: safety in intimate relationships, comfort with physical contact from parents or primary caregivers, ease of using public restrooms, general bathroom comfort, bodily safety, and enjoyment of consensual sexual activities. Respondents rated their agreement with related statements on a 0 (completely uncomfortable) to 100 (completely comfortable) sliding scale.

#### Engagement in risky behaviors

To assess the frequency of respondents’ engagement in behaviors that could potentially compromise their health or safety, we evaluated the association of various lifestyle variables with VCUG exposure. The assessed behaviors included regular smoking, beyond regular alcohol consumption, regular physical inactivity, regular work absenteeism, and regular drug use.

Branching logic was applied to questions relevant only to female participants (gynecological visits, giving birth, pap smears, etc.). We asked about VCUG exposure only at the end of the main survey component to avoid influencing respondents’ prior answers related to trauma and health behaviors. We also collected data on Adverse Childhood Experiences (ACEs) to investigate potential influences on respondent answers. For the purposes of this study, we will not consider ACES or other factors in our analyses.

### STATISTICAL APPROACH

Data management was performed using R and Microsoft Excel. Descriptive statistics were used to summarize standard demographic and baseline characteristics of the study population.

SAS/FREQ or RStudio were used to perform statistical analyses. We used T-tests with necessary corrections to determine variance when comparing continuous variables between the VCUG and control groups, and chi-square tests to examine any associations between categorical variables and VCUG status. We used Fisher’s exact test to analyze variables with insubstantial data.

## Results

### RESPONDENTS

A total of 399 respondents participated in the study. 334 respondents answered all required survey sections in the main survey component and thus were included in the study. We identified a total of 204 respondents (61%) in the VCUG group, and a total of 130 respondents in the Control group (39%). Demographic characteristics of the two groups are shown in Table 1A.

**TABLE 1A.**
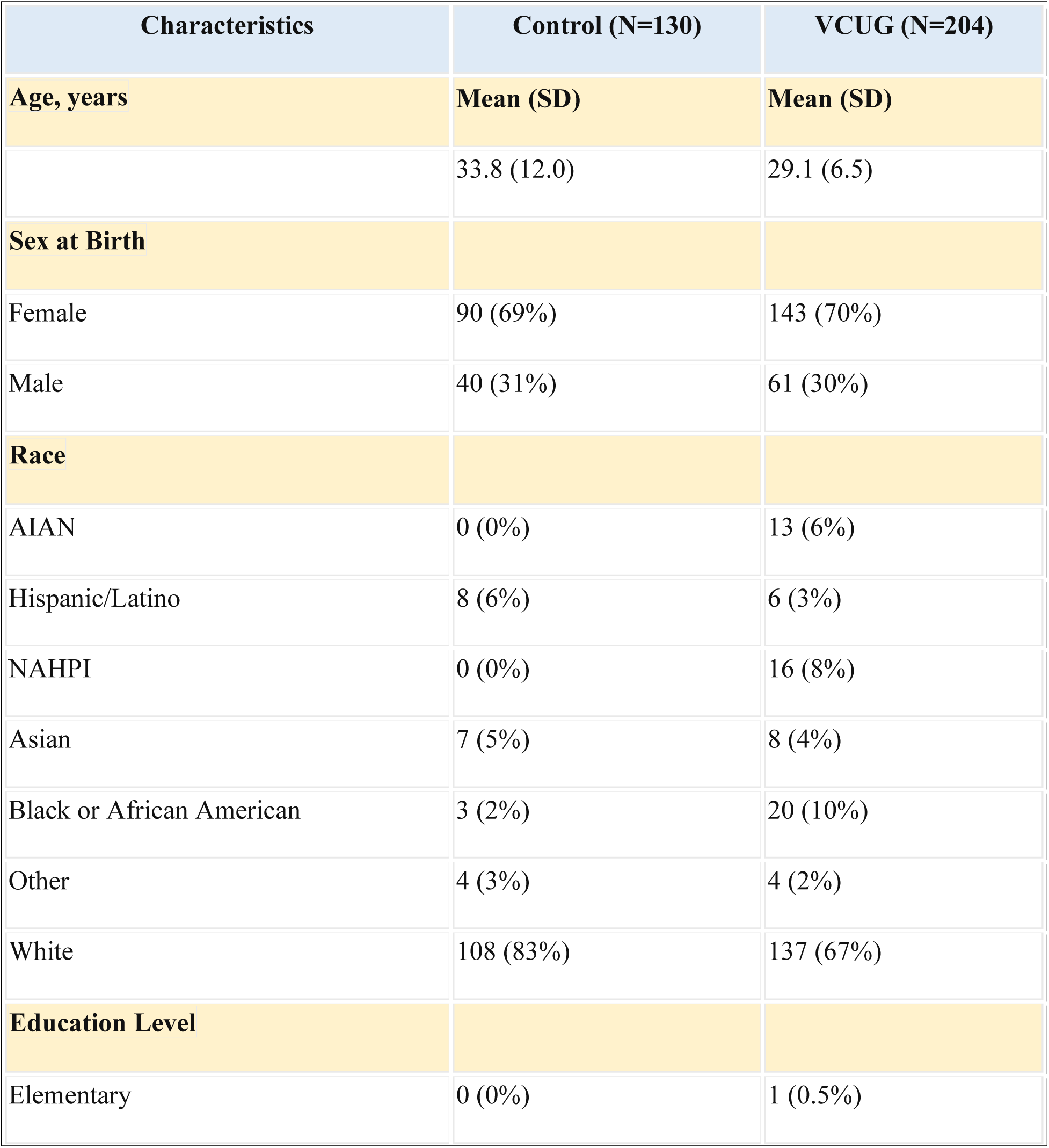
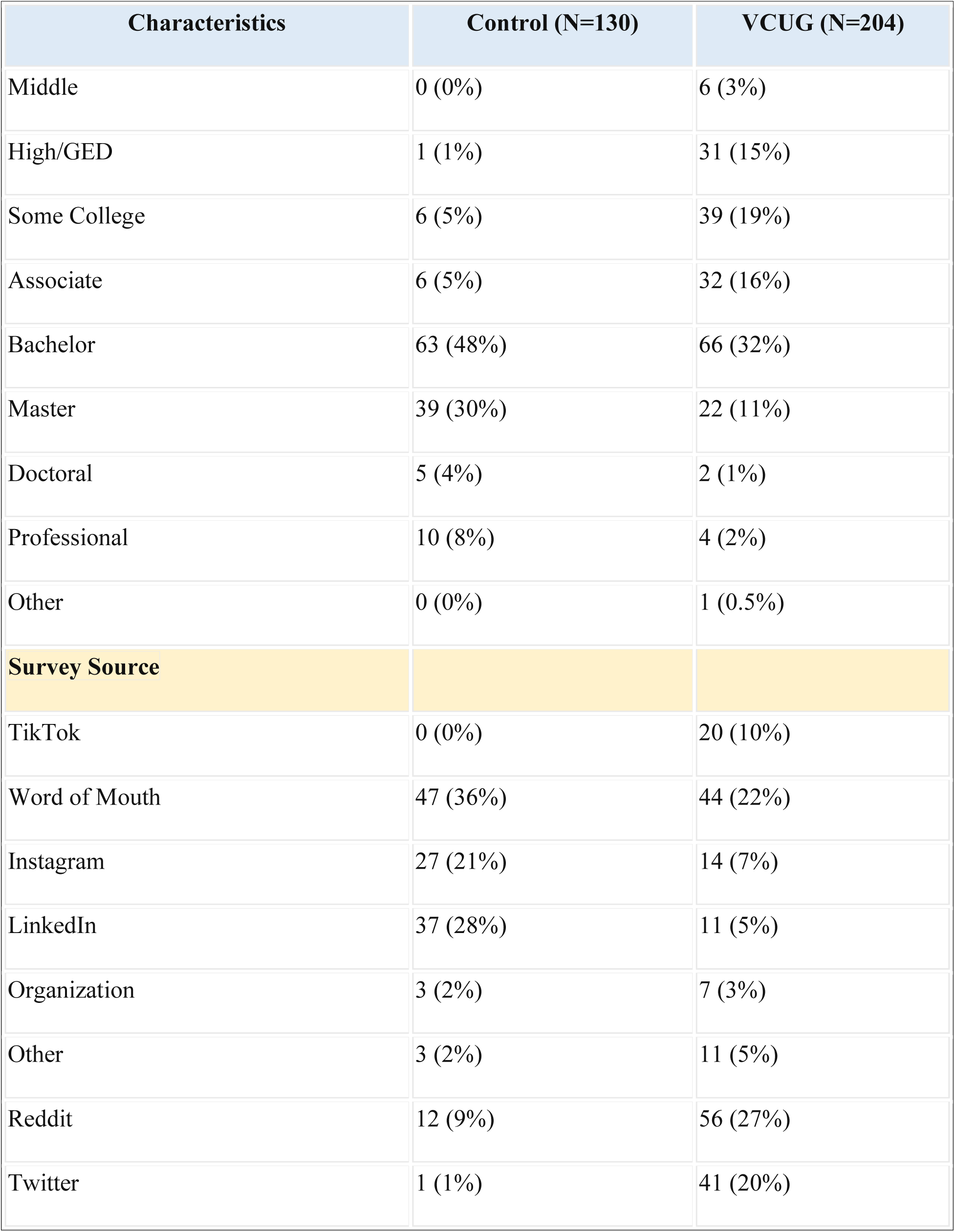
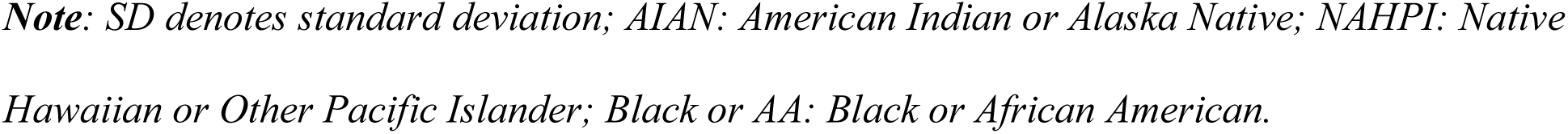
DEMOGRAPHIC AND BASELINE RESPONDENT CHARACTERISTICS.

| Characteristics | Control (N=130) | VCUG (N=204) |
| --- | --- | --- |
| **Age, years** | **Mean (SD)** | **Mean (SD)** |
|  | 33.8 (12.0) | 29.1 (6.5) |
| **Sex at Birth** |  |  |
| Female | 90 (69%) | 143 (70%) |
| Male | 40 (31%) | 61 (30%) |
| **Race** |  |  |
| AIAN | 0 (0%) | 13 (6%) |
| Hispanic/Latino | 8 (6%) | 6 (3%) |
| NAHPI | 0 (0%) | 16 (8%) |
| Asian | 7 (5%) | 8 (4%) |
| Black or African American | 3 (2%) | 20 (10%) |
| Other | 4 (3%) | 4 (2%) |
| White | 108 (83%) | 137 (67%) |
| **Education Level** |  |  |
| Elementary | 0 (0%) | 1 (0.5%) |
| Middle | 0 (0%) | 6 (3%) |
| High/GED | 1 (1%) | 31 (15%) |
| Some College | 6 (5%) | 39 (19%) |
| Associate | 6 (5%) | 32 (16%) |
| Bachelor | 63 (48%) | 66 (32%) |
| Master | 39 (30%) | 22 (11%) |
| Doctoral | 5 (4%) | 2 (1%) |
| Professional | 10 (8%) | 4 (2%) |
| Other | 0 (0%) | 1 (0.5%) |
| **Survey Source** |  |  |
| TikTok | 0 (0%) | 20 (10%) |
| Word of Mouth | 47 (36%) | 44 (22%) |
| Instagram | 27 (21%) | 14 (7%) |
| LinkedIn | 37 (28%) | 11 (5%) |
| Organization | 3 (2%) | 7 (3%) |
| Other | 3 (2%) | 11 (5%) |
| Reddit | 12 (9%) | 56 (27%) |
| Twitter | 1 (1%) | 41 (20%) |

*Note: SD denotes standard deviation; AIAN: American Indian or Alaska Native; NAHPI: Native Hawaiian or Other Pacific Islander; Black or AA: Black or African American.*

**TABLE 1B.**
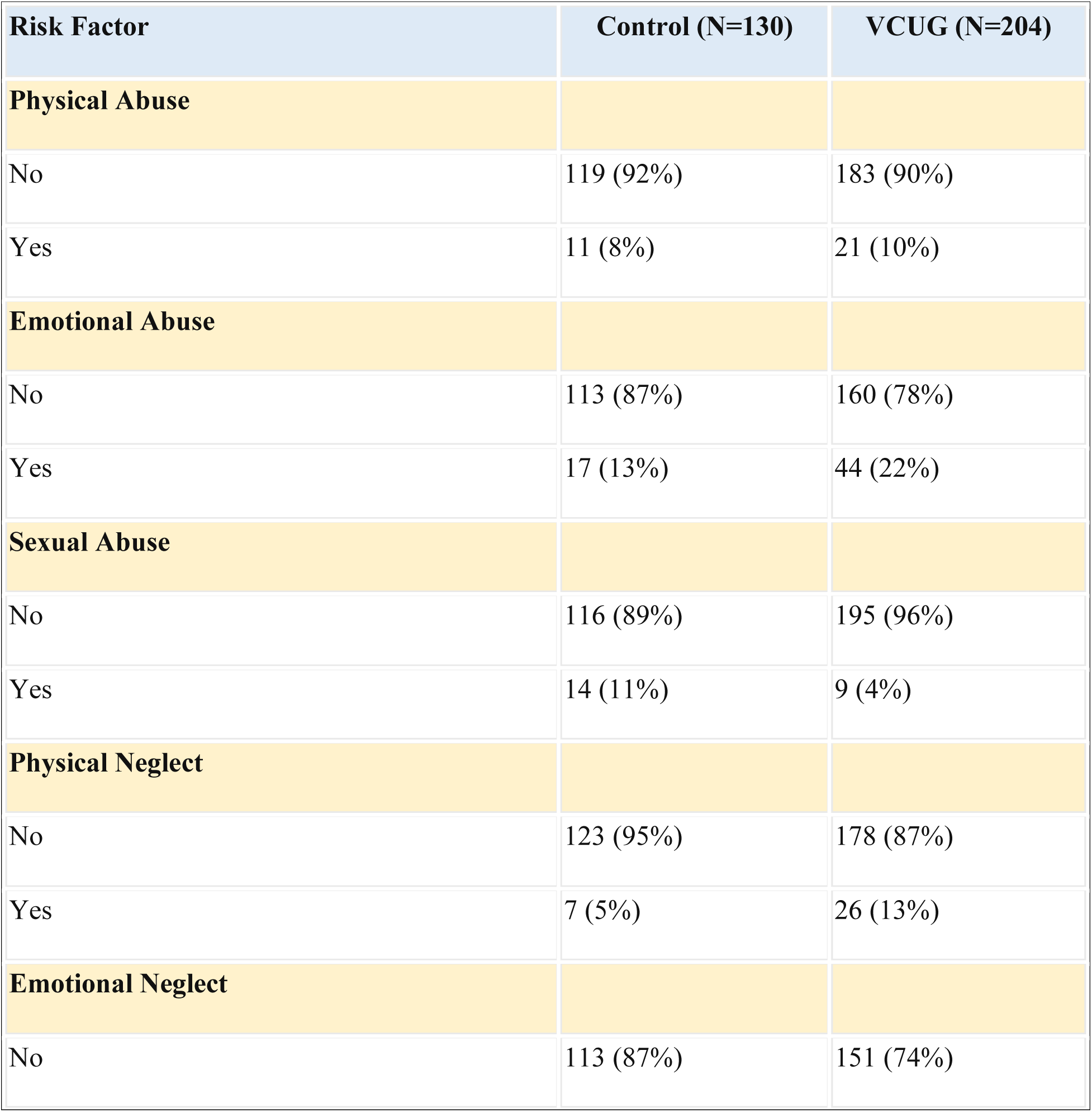
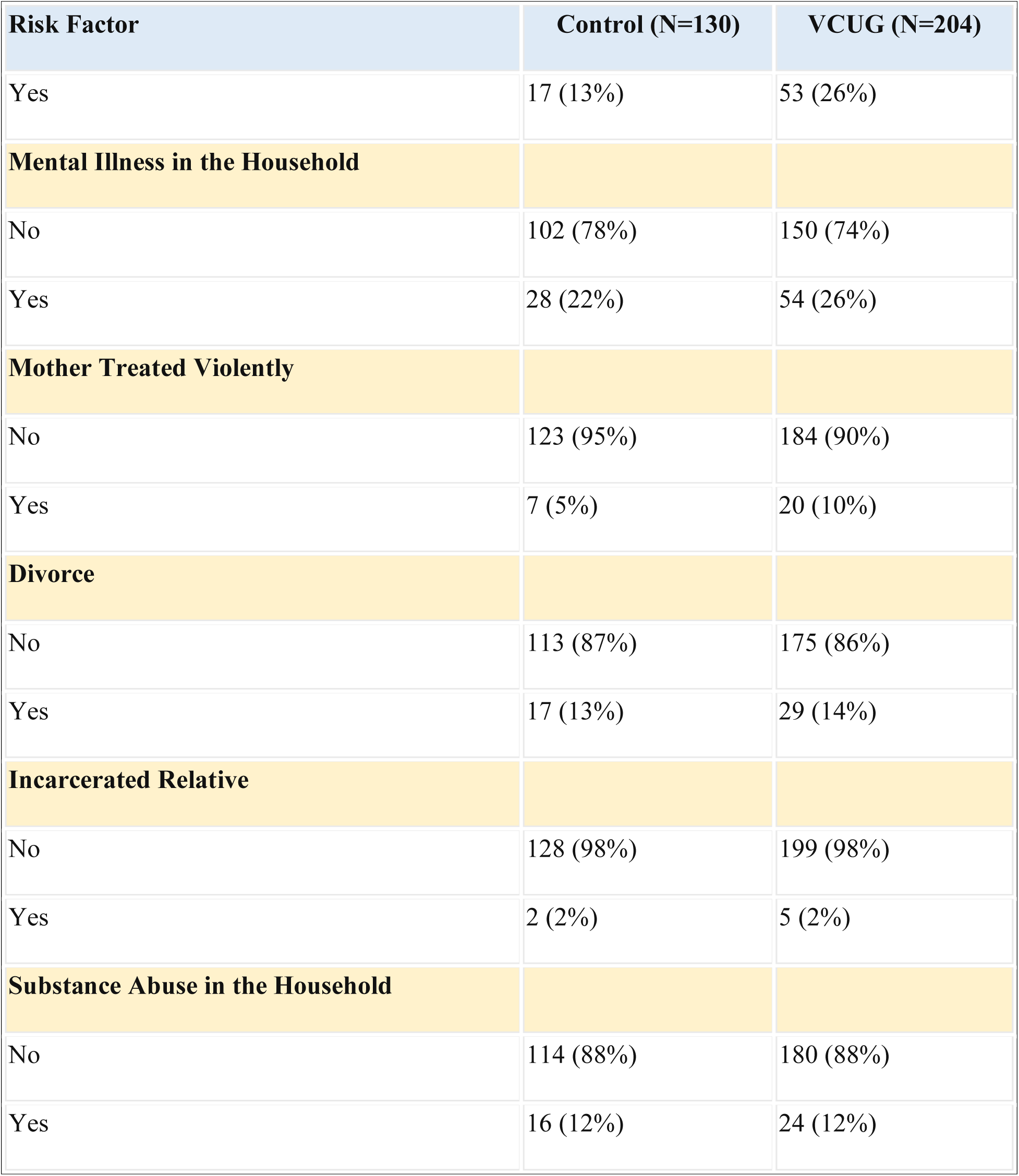
SUMMARY OF DISTRIBUTION OF POTENTIAL CONFOUNDERS FOR TRAUMA: PREVALENCE OF ADVERSE CHILDHOOD EXPERIENCES (ACES)

### HEALTH OUTCOMES

The study revealed significant disparities in physical and mental health perceptions between the VCUG group and the control group. Participants in the VCUG group reported markedly lower mean scores for physical health (M=63.43, SD=22.46) compared to the control group (M=72.58, SD=18.85), with a 95% confidence interval (diff 4.66-13.64, p<0.001). Similarly, the VCUG group reported significantly lower mean scores for mental health (M=54.63, SD=24.27) than controls (M=66.70, SD=19.30), with a 95% confidence interval (diff 7.35-16.79, p<0.001).

These findings underscore a potentially substantial impact of VCUG exposure on both physical and mental health. Detailed distributive and statistical analyses can be found in Table 2.

**TABLE 2.** SUMMARY OF RESULTS FOR HEALTH PERCEPTIONS.

| Health Aspect | Group | N | Mean (M) | Standard Deviation (SD) | T-value (t) | P-value (p) | Confidence Interval (CI) |
| --- | --- | --- | --- | --- | --- | --- | --- |
| Physical Health Rating | VCUG | 204 | 63.43 | 22.46 | 307.85 | <0.001 | [4.66, 13.64] |
|  | Control | 130 | 72.58 | 18.85 |  |  |  |
| Mental Health Rating | VCUG | 204 | 54.63 | 24.27 | 316.03 | <0.001 | [7.35, 16.79] |
|  | Control | 130 | 66.70 | 19.30 |  |  |  |

Further examination of the prevalence of specific illnesses and psychological conditions among the VCUG group revealed concerning trends. Anxiety diagnoses were reported by 56% of the VCUG group, compared to 35% of controls, with a chi-square test confirming a significant association (χ²=14.40, p=0.0001). Similarly, depression diagnoses were higher in the VCUG group (47%) compared to controls (27%), with significant association (χ²=13.51, p=0.0002).

PTSD diagnoses were also more prevalent in the VCUG group (25%) compared to controls (12%), confirmed by chi-square test (χ²=9.64, p=0.002). For OCD, 13% of the VCUG group reported diagnoses compared to 5% of controls, with a significant association (χ²=6.63, p=0.01).

An analysis of RGS/PGAD diagnoses indicated that 1% of the VCUG group reported this condition compared to none in the control group. Due to the low expected counts, Fisher’s exact test was employed, confirming no significant association (p=0.29). However, 15% of the VCUG group indicated that their symptoms might suggest RGS/PGAD compared to 2% of controls, with chi-square tests revealing a significant association when including “maybe” respondents (χ²=18.12, p<0.001). Similar trends were observed for HPF, with 16% of the VCUG group reporting diagnoses versus 2% of controls (χ²=15.15, p<0.001), and 28% indicating likely HPF symptoms compared to 5% of controls (χ²=43.35, p<0.001). For BPD, 9% of the VCUG group reported diagnoses compared to 4% of controls, with no significant association found (χ²=3.07, p=0.08). Detailed analyses are provided in Table 3.

**TABLE 3.** SUMMARY OF RESULTS FOR LONG-TERM ILLNESSES, IN ORDER OF STRENGTH OF ASSOCIATION TO VCUG EXPOSURE.

| Condition | Chi-Square ($\chi^2$) | DF | P-value | VCUG Diagnosed (N, %) | Control Diagnosed (N, %) |
| --- | --- | --- | --- | --- | --- |
| Anxiety | 14.40 | 1 | 0.0001 | 114 (56%) | 46 (35%) |
| Depression | 13.51 | 1 | 0.0002 | 96 (47%) | 35 (27%) |
| Post-traumatic stress disorder (PTSD) | 9.64 | 1 | 0.002 | 51 (25%) | 16 (12%) |
| Obsessive-compulsive disorder (OCD) | 6.63 | 1 | 0.01 | 27 (13%) | 6 (5%) |
| Bipolar disorder (BPD) | 3.07 | 1 | 0.08 | 18 (9%) | 5 (4%) |
| Restless genital syndrome (RGS) or persistent genital arousal disorder (PGAD) | Fisher's | 1 | 0.29 | 2 (1%) | 0 (0%) |
| Hypertonic pelvic floor (HPF) | 15.50 | 1 | <0.001 | 33 (16%) | 3 (2%) |

Regarding medical care refusal, significant differences were noted among the VCUG group - particularly female-born participants. 20% of female-born VCUG respondents report complete refusal of pap smears, compared to 1% of the female control group, with a significant association (χ²=17.29, p<0.001). Similarly, 15% of the female VCUG cohort reported completely refusing gynecological care compared to 2% of female controls (χ²=10.36, p=0.001). In both men and women, 11% of VCUG respondents completely refuse urological care compared to 5% of controls (χ²=4.44, p=0.04). Complete refusal of general physician care was reported by 3% of the VCUG group compared to 2% of controls, with Fisher’s exact test indicating no significant association (p=0.49). Refer to Table 4 for comprehensive analyses.

**TABLE 4.**
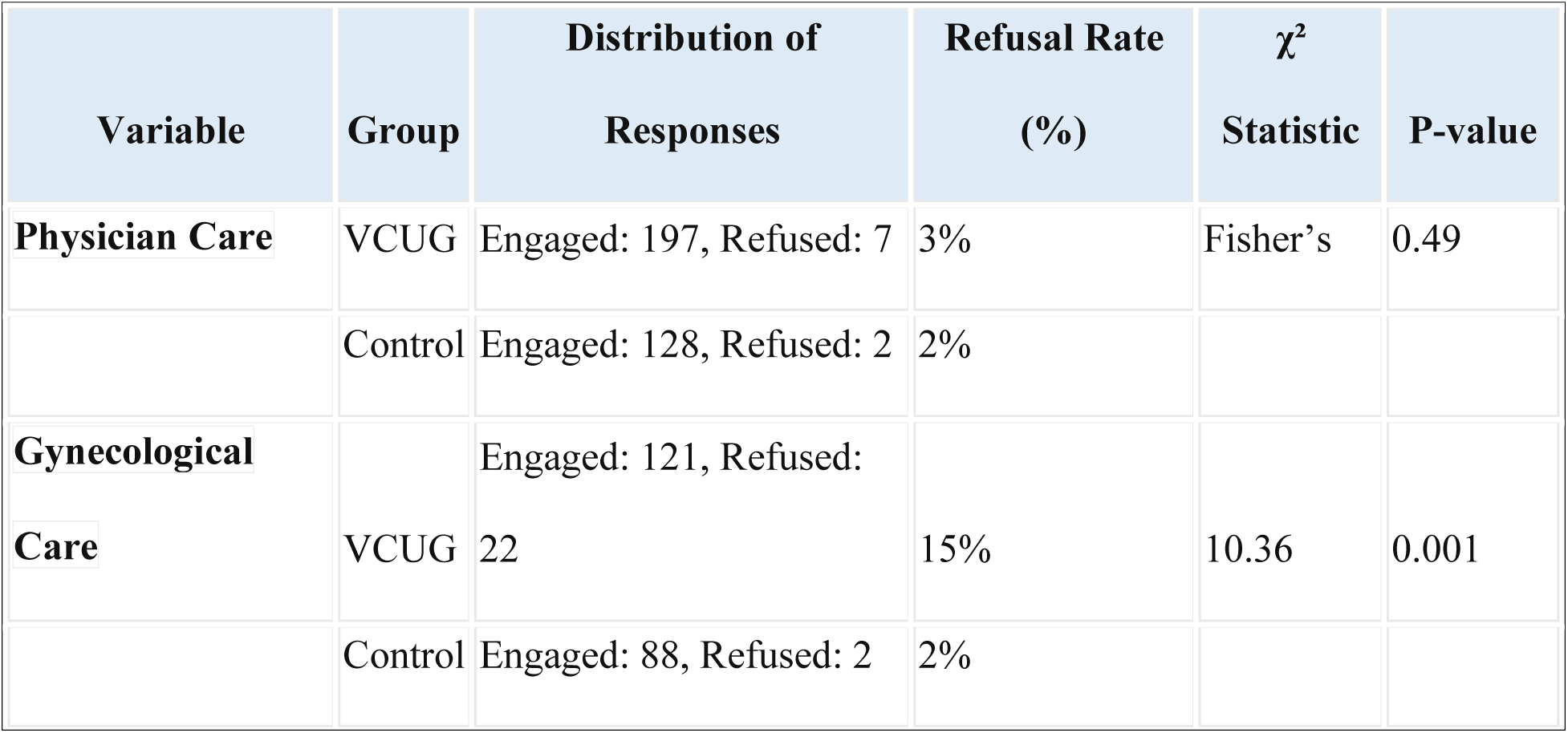
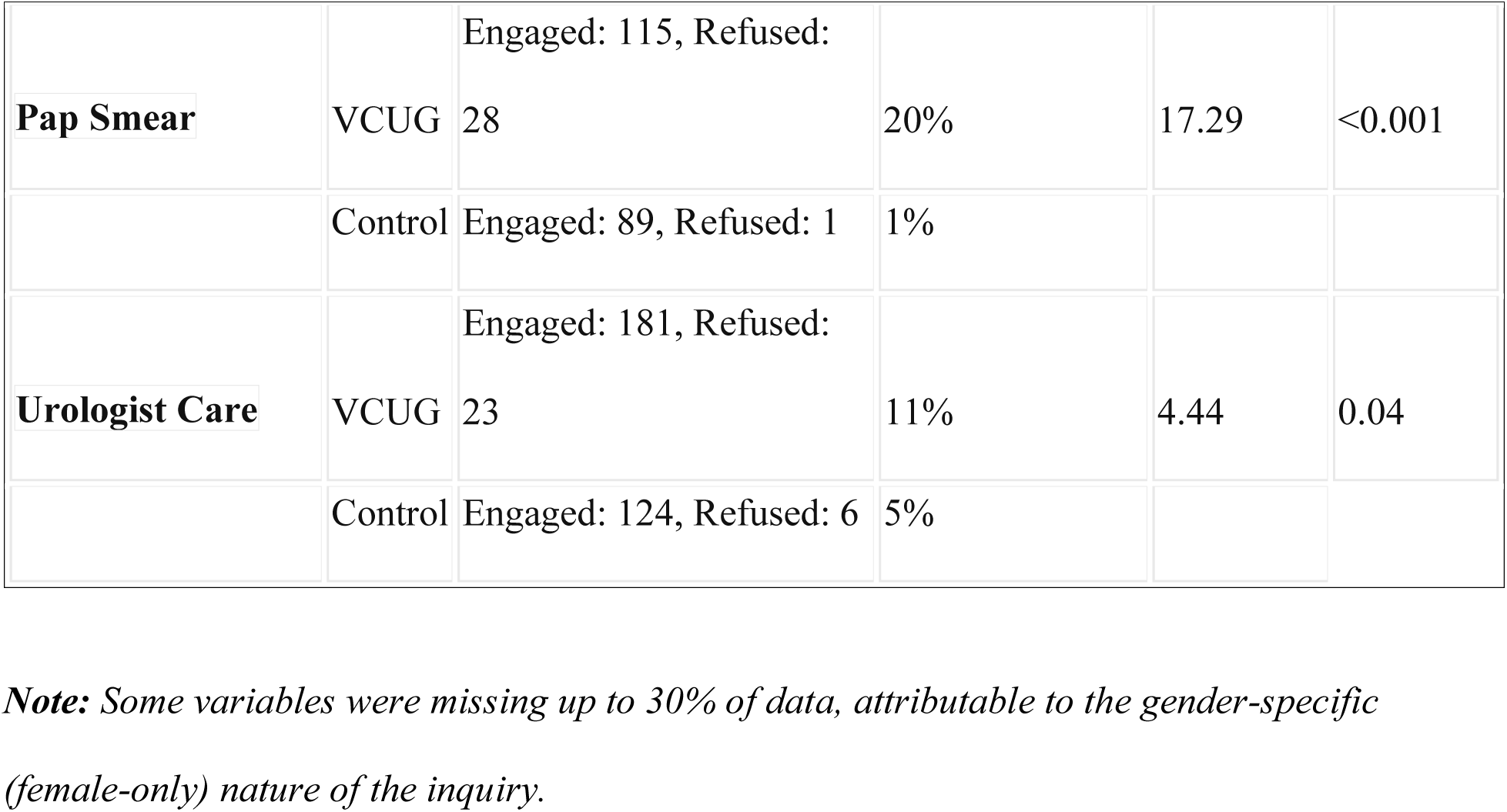
SUMMARY OF RESULTS FOR ATTITUDES TOWARDS VARIOUS TYPES OF MEDICAL CARE.

Quality of life factors also showed significant differences. The VCUG group reported lower scores in every quality of life area we asked about: safety in intimate relationships (M=67.22, SD=26.26 vs. M=84.89, SD=21.56; 95% CI of diff 12.49-22.87, p<.001), physical contact from parents/caregivers (M=56.77, SD=28.68 vs. M=87.15, SD=20.71; 95% CI of diff 25.06-35.71, p<.001), using public restrooms (M=67.91, SD=28.69 vs. M=86.67, SD=21.24; 95% CI of diff 13.37-24.15, p<.001), general restroom use (M=71.39, SD=25.85 vs. M=92.17, SD=16.01; 95% CI of diff 16.27-25.28, p<.001), feeling safe in one’s body (M=60.23, SD=28.67 vs. M=83.97, SD=24.50; 95% CI of diff 17.75-29.72, p<.001), and enjoyment of consensual sexual activities (M=61.72, SD=29.66 vs. M=84.68, SD=23.13; 95% CI of diff 17.20-28.72, p<.001). Complete analyses can be found in Table 5.

**TABLE 5.**
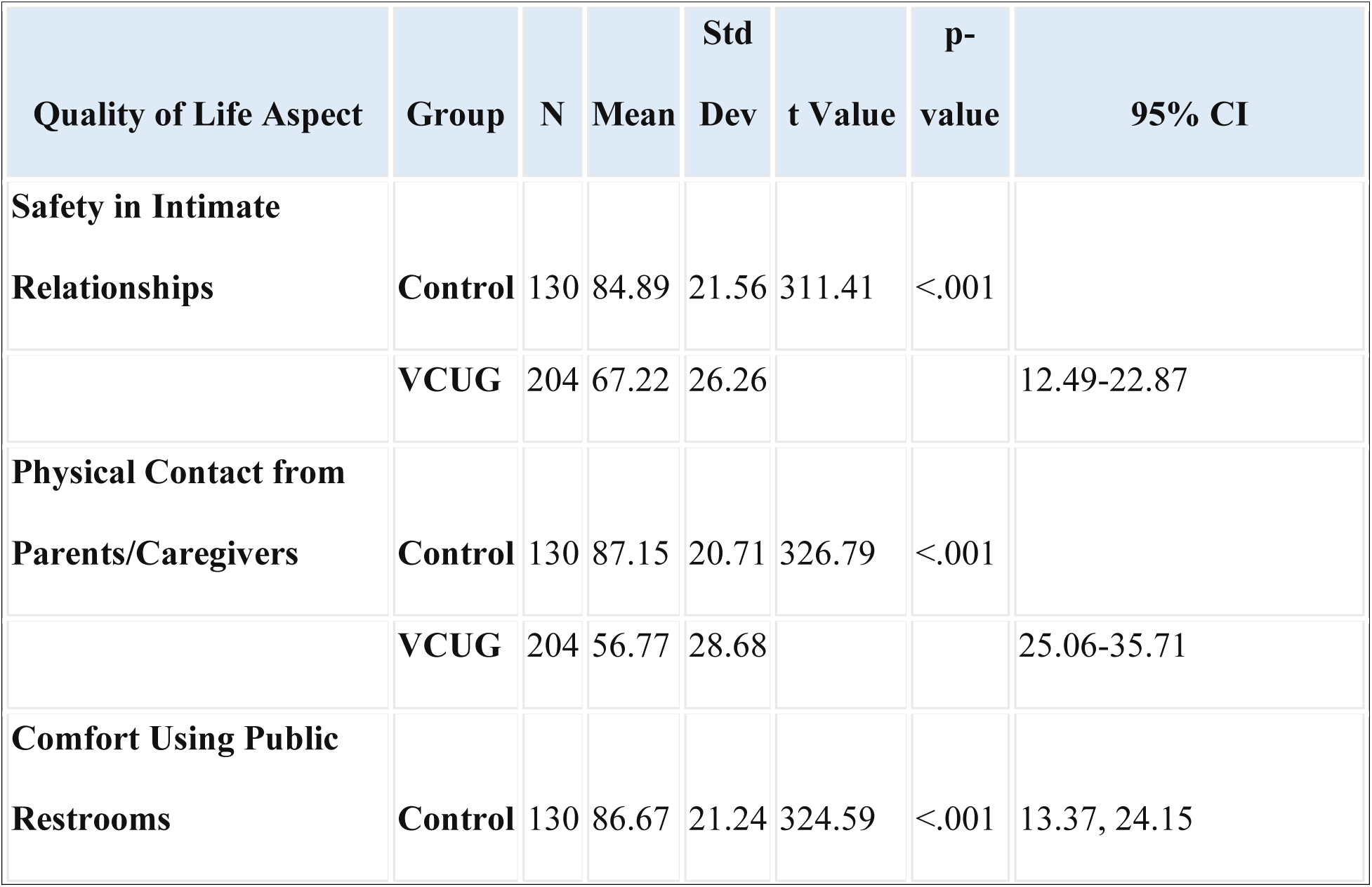
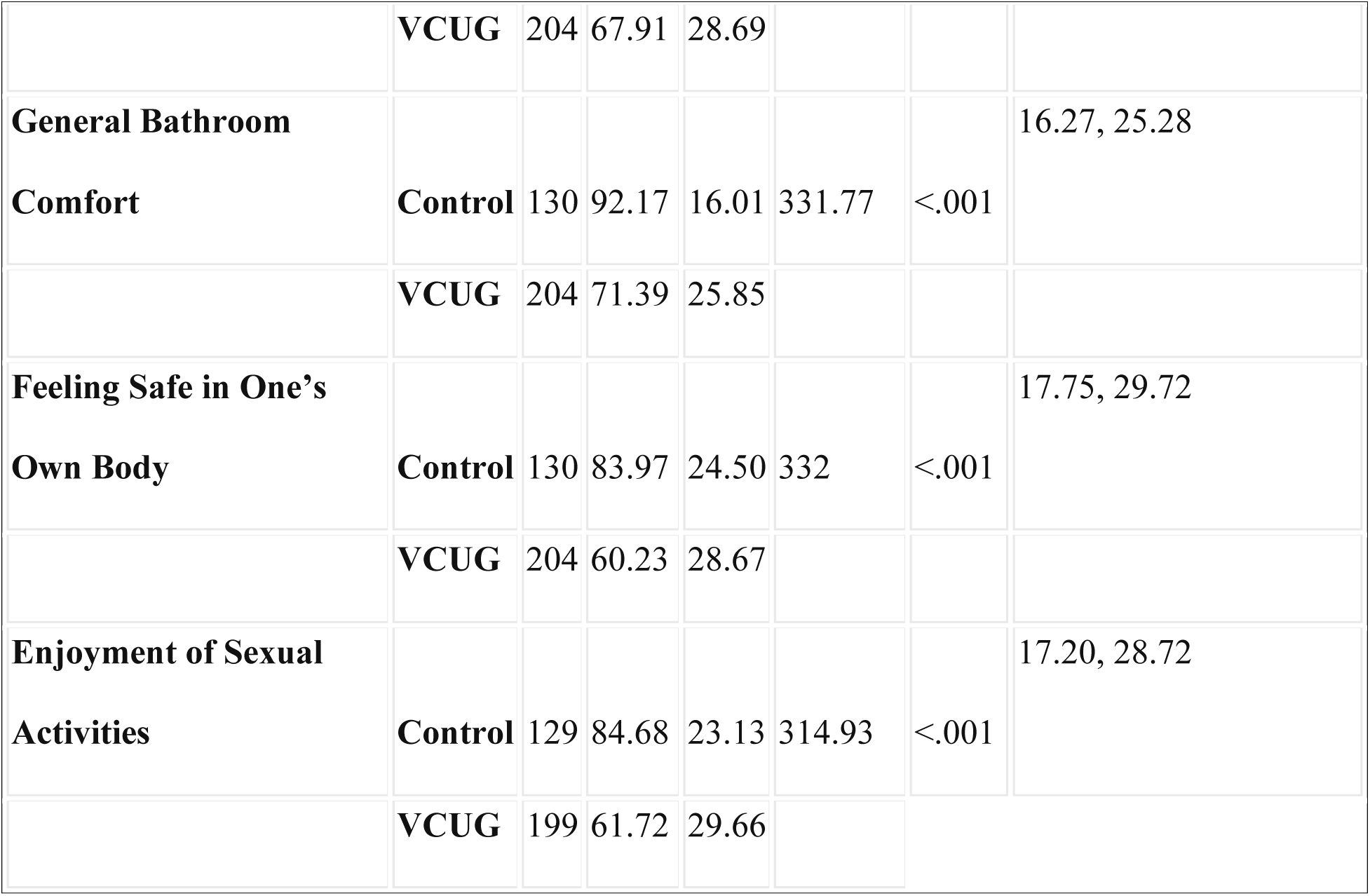
SUMMARY OF RESULTS FOR ADULT QUALITY OF LIFE BEHAVIORS.

Lastly, the study explored risky behaviors, revealing significant associations between VCUG status and regular smoking (χ²=35.85, p<0.001; 34% of VCUG group vs. 5% of controls), missing work (χ²=11.61, p=0.0006; 11% of VCUG group vs. 1% of controls), excessive alcohol use (χ²=6.74, p=0.009; 21% of VCUG group vs. 9% of controls), and lack of physical activity (χ²=9.33, p=0.002; 28% of VCUG group vs. 13% of controls). No significant association was found between VCUG status and regular drug use (χ²=2.56, p=0.61), though 11% of the VCUG group reported such behavior compared to 8% of controls. For a detailed breakdown, refer to Table 6.

**TABLE 6.** SUMMARY OF RESULTS FOR RISKY BEHAVIORS.

| **Lifestyle Behavior** | **Control Group %**<br>**"Yes" (n=130)** | **VCUG Group %**<br>**"Yes" (n=204)** | **Chi-Square**<br>**(X<sup>2</sup>)** | **p-value** |
| --- | --- | --- | --- | --- |
| **Regular lack of**<br>**physical activity** | 13% | 28% | 9.33 | 0.002 |
| **Regularly missing**<br>**work** | 1% | 11% | 11.61 | 0.0007 |
| **Beyond regular**<br>**alcohol use** | 9% | 21% | 6.74 | 0.009 |

| Lifestyle Behavior | Control Group %<br>“Yes” (n=130) | VCUG Group %<br>“Yes” (n=204) | Chi-Square<br>(X<sup>2</sup>) | p-value |
| --- | --- | --- | --- | --- |
| Regular smoking | 5% | 34% | 35.85 | <0.001 |
| Regular drug use | 8% | 11% | 0.26 | 0.61 |

**TABLE 7.**
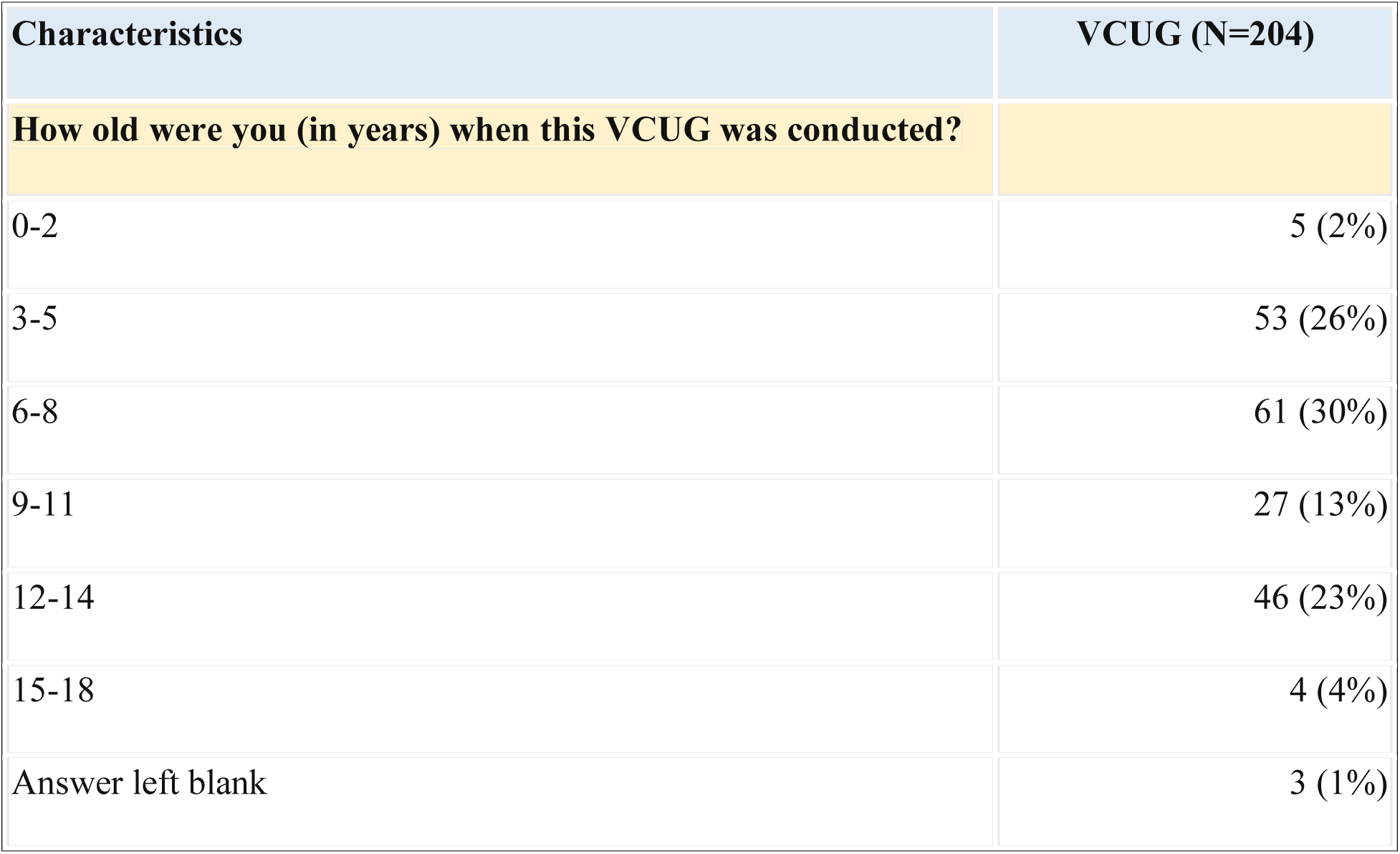
DISTRIBUTION OF AGE IN YEARS AT TIME OF FIRST OR MOST MEMORABLE VCUG (VCUG GROUP ONLY)

**TABLE 8.** DISTRIBUTION OF NUMBER OF VCUGS CONDUCTED PER PATIENT.

| Characteristics | VCUG (N=204) |
| --- | --- |
| **How many VCUGs do you remember experiencing before the age of 18?** |  |
| 1 | 117 (57%) |
| 2 | 41 (20%) |
| 3 | 24 (12%) |
| 4 | 8 (4%) |
| 5 | 1 (0%) |
| 6 | 4 (2%) |
| 7 | 2 (1%) |
| 8 | 3 (1%) |
| 9 | 0 (0%) |
| 10 | 1 (0%) |
| Answer left blank | 3 (1%) |

**TABLE 9.**
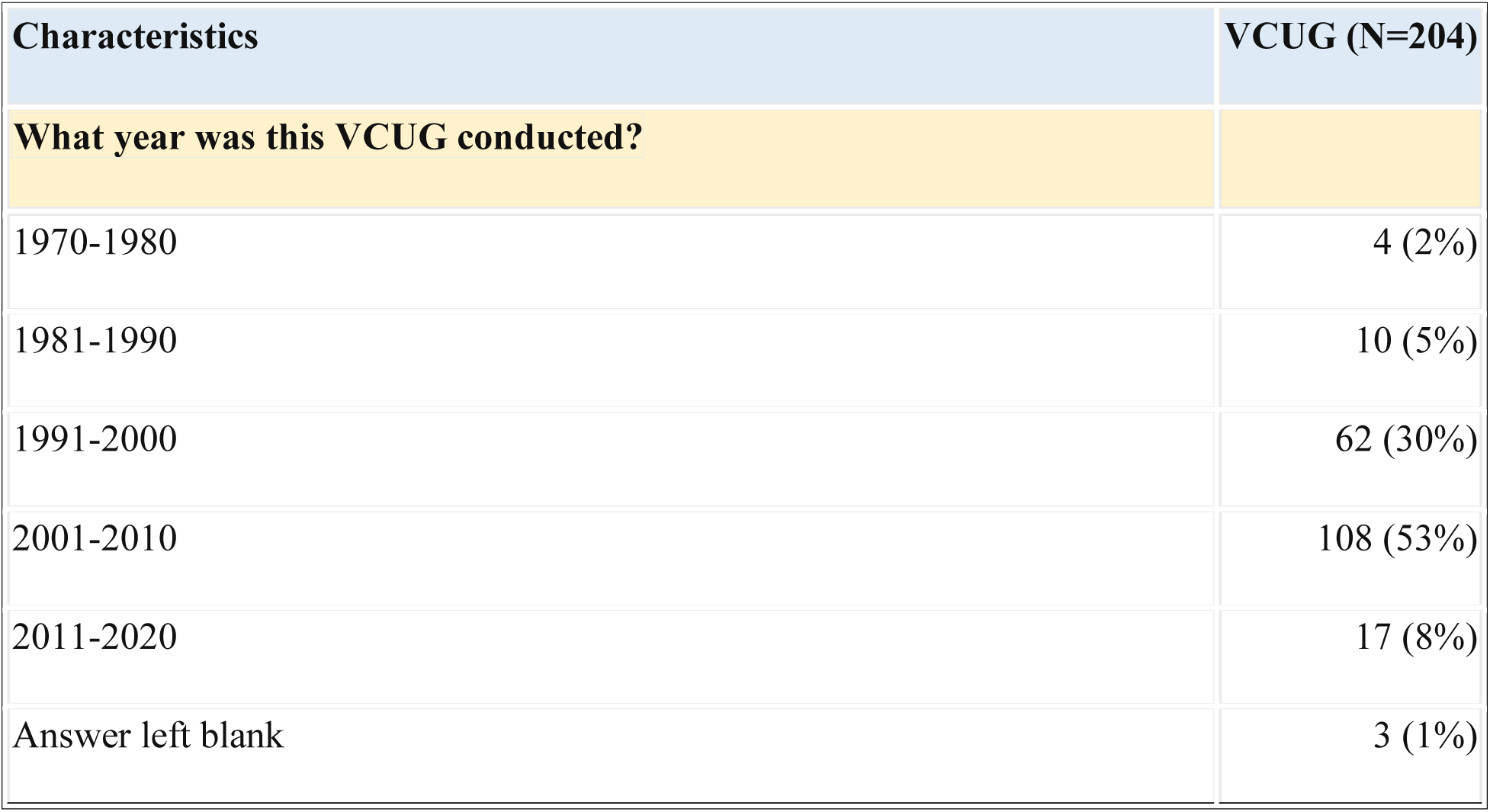
DISTRIBUTION OF YEAR AT TIME OF FIRST OR MOST MEMORABLE VCUG.

| Characteristics | VCUG (N=204) |
| --- | --- |
| **What year was this VCUG conducted?** |  |
| 1970-1980 | 4 (2%) |
| 1981-1990 | 10 (5%) |
| 1991-2000 | 62 (30%) |
| 2001-2010 | 108 (53%) |
| 2011-2020 | 17 (8%) |
| Answer left blank | 3 (1%) |

**TABLE 10.** STATE IN WHICH FIRST OR MOST MEMORABLE VCUG TOOK PLACE, IN ORDER OF FREQUENCY. “If you do not remember your first VCUG, please use the VCUG most available in your memory to answer the following section.”

| Characteristics | VCUG (N=204) |
| --- | --- |
| **In what state was this VCUG conducted?** |  |
| California | 28 (14%) |
| Massachusetts | 14 (7%) |
| Illinois | 13 (6%) |
| New York | 12 (6%) |
| Florida | 10 (5%) |
| Hawaii | 10 (5%) |
| New Jersey | 8 (4%) |
| Indiana | 8 (4%) |
| Texas | 7 (3%) |
| Ohio | 6 (3%) |
| Virginia | 6 (3%) |
| Washington | 6 (3%) |
| Minnesota | 6 (3%) |
| Michigan | 6 (3%) |
| Alabama | 5 (2%) |
| Colorado | 4 (2%) |
| Wisconsin | 4 (2%) |
| Georgia | 4 (2%) |
| Pennsylvania | 4 (2%) |
| Iowa | 3 (1%) |
| Arizona | 3 (1%) |
| Maryland | 3 (1%) |
| Missouri | 3 (1%) |
| Nevada | 3 (1%) |
| Utah | 3 (1%) |
| Arkansas | 3 (1%) |
| Connecticut | 3 (1%) |
| Idaho | 1 (0%) |
| Nebraska | 1 (0%) |
| Kansas | 1 (0%) |
| Oklahoma | 1 (0%) |
| North Carolina | 1 (0%) |
| New Mexico | 1 (0%) |
| Delaware | 1 (0%) |
| South Carolina | 1 (0%) |
| West Virginia | 1 (0%) |
| Tennessee | 1 (0%) |
| Wyoming | 1 (0%) |
| Kentucky | 1 (0%) |
| New Hampshire | 1 (0%) |
| Maine | 1 (0%) |
| Vermont | 1 (0%) |
| All other states | 0 (0%) |
| Answer left blank | 8 (4%) |

## Discussion

This exploratory retrospective cohort study illuminates the risk of long-standing repercussions of childhood VCUG on patient health and well-being, encompassing both physical and psychological domains.

Individuals with a history of VCUG report markedly poorer perceptions of their physical and mental health compared to controls. These disparities in health perception are substantiated by more individuals in the VCUG cohort reporting certain chronic conditions and illnesses.

The observed refusal of the VCUG cohort, particularly among women, to seek routine and specialized medical care, highlights a profound response to the procedure. Avoiding preventive healthcare not only endangers individual health outcomes by elevating the risk of late diagnoses, but can also contribute to rising healthcare costs associated with treating advanced conditions such as cancer. Considering that VCUG patients may be less likely to seek medical care in general, these rates may be underreported.

We recognize the potential recall and sample bias inherent in this study, attributable to legitimate challenges encountered during the study’s execution due to lack of institutional resources. In respondents, the process of accurately recalling or identifying personal VCUG history may be compromised regardless of trauma status. Among the respondents who *do* remember, individuals with pronounced reactions to their experiences are disproportionately represented on social media platforms such as Reddit. Consequently, 27% of the VCUG cohort found our study through Reddit compared to 9% of the control group (see Table 1A). Further research could build upon the foundation we have laid here and seek to better control for potential confounders.

Despite these limitations, our study highlights an urgent call to review VCUG informed consent protocols, clinical practice and ordering guidelines, and informational materials to patients, families, and clinicians.

This study suggests that the impact of VCUG might extend years beyond the immediate procedure and into adulthood. The preliminary insights from this study should encourage a broader discussion on the application of VCUG, ensuring that the risk of trauma is weighed against clinical benefits. Despite being a standard diagnostic tool, VCUG’s potentially pervasive and long-lasting psychological effects necessitate a call for a more thorough risk assessment by healthcare institutions. The need for age-appropriate informed consent and trauma-informed care practices is imperative - patients and guardians are often unaware of these potential outcomes, which should be integral to the consent dialogue. This should be paired with enforced protocols from guideline-setting pediatric institutions. Finally, these findings indicate a clear need for a more rigorous exploration of VCUG alternatives that minimize psychological trauma. There is a critical gap in non-invasive yet equally effective diagnostic methods for reflux. Such innovation would represent a substantial advancement in pediatric urological care, patient advocacy, and reduced future burden on the healthcare system overall.

## Data Availability

All data produced in the present study are available upon reasonable request to the authors

## Acknowledgements

We extend our sincere gratitude to Scott Appel and the Biostatistics Analysis Center at the University of Pennsylvania for their invaluable guidance and statistical analysis throughout this study.

